# Quality of Counselling, Exposure to Vaccination Messages and Caregivers’ Knowledge on the Uptake of Penta Vaccine in Six Northern Nigerian States

**DOI:** 10.1101/2024.08.09.24311716

**Authors:** Matthew Alabi, Leanne Dougherty, Eno-Obong Etim, Adebola Adedimeji

## Abstract

**Background:** Pentavalent 3 (Penta-3) coverage for children aged 12-23 months is used as the prime substitute for determining vaccination coverage and monitoring the performance of the national immunization programme. However, the coverage for Penta 3 in Nigeria remains low. Quality interaction between caregivers and providers and access to appropriate information are associated with healthcare utilization and acceptance of recommended health behaviours. This study examines the quality of counselling (QOC), caregivers’ exposure to vaccination messages and child’s uptake of the Penta vaccine.

**Methods:** This was a cross-sectional study that utilized quantitative data obtained through a survey. Caregivers (n=561) of children aged 2-24 months accessing child vaccination services who received Penta vaccines at randomly selected health facilities (n=163) offering routine immunization (RI) services were surveyed. Penta uptake was coded as binary; hence, binary logistic regression was performed using Stata 14.

**Result:** We found that 56% of the caregivers received quality counselling. Although awareness of child vaccination was high (70%), two-thirds had poor exposure. The uptake of all three doses of Penta vaccine was 43%. Contextual factors associated with Penta uptake include caregivers’ knowledge of when a child should receive their first vaccination (aOR=2.08; 95% CI=1.01-4.29), sources of child vaccination messages, namely, place of worship (aOR=2.78; 95% CI=1.15-6.67), Community Health Workers CHW (aOR=1.95; 95% CI=1.14-3.34), community leader (aOR=2.21; 95% CI=1.11-4.41) and residence in the northwest region (aOR=2.60; 95% CI=1.51-4.48).

**Conclusion:** Given the low quality of counselling and the positive influence of religious and traditional leaders, interventions that prioritize strengthening patient-provider interaction and community structure are crucial for increasing child vaccination coverage in Nigeria.

## BACKGROUND

Childhood immunization during the first few months of life seeks to build immunity and is needed for protection against infectious diseases during infancy^1^. The routine child immunization schedule recommends that a child receive a specific vaccine at a particular age. According to the World Health Organization (WHO), the childhood vaccination program aims to ensure that children receive appropriate doses of vaccine at a particular age without delay ^2^. While full-child vaccination coverage is considered a better indicator for measuring the complete benefits of childhood immunization^3^, Penta 3 coverage for children aged 12-23 months has been frequently used as the prime substitute for determining vaccination coverage and monitoring the performance of national immunization programmes^4^.

In Nigeria, the coverage for the Penta vaccine and all the recommended doses of routine immunization has remained low, far below the targets set by WHO in the 2012 Global Vaccine Action Plan for ensuring universal access to immunization^5,6^. This is important as donors often consider the coverage for Penta and measles vaccines because they constitute indicators of health system performance ^7,8^ and because of the inclusion of Penta in the original expanded program on immunization (EPI) due to its multidose nature ^9^. The Pentavalent vaccine combines five vaccines in one, administered to protect against life-threatening diseases – Diphtheria, Tetanus, whooping cough, Hepatitis B, and Haemophilus influenza type B, all contained in a single dose^1^. However, in 2019, Nigeria had the highest number of infants unvaccinated with the Penta-1 vaccine through routine immunization services ^10^. Additionally, while the global coverage for Penta-3 was estimated at 81% in 2021^11^, the coverage for Penta-3 in Nigeria as of 2021 remains very low at 56% when compared to that of neighbouring countries such as Ghana (98%), Senegal (85%) and Niger (82%)^12^. Data from Nigeria suggest regional differences in Penta-3 coverage, with rates as low as 19% in the Northeast and 52% in the Southwest ^13^, relative to the higher coverage reported in other regions.

Studies^14–21^ have identified the influence of individual (age, level of education, place of residence), health-seeking behaviour factors (antenatal care attendance, postnatal care attendance, facility delivery), and community-level factors such as distance to a health facility and the role of traditional and institutional structure on routine child immunization uptake in Nigeria ^16,18,22,23^. These studies reported significant effects of maternal characteristics and health-seeking behaviour factors, and also acknowledged the important role of traditional and religious institutions in disseminating key health information, including routine child immunization uptake, due to their strong influence on their subjects. Disseminating immunization messages using coordinated community structures that convey targeted information via different channels, including town criers and the use of short messaging services (SMS), were evidence of strong and effective communication strategies in addition to being culturally acceptable.

However, research into the importance of service delivery factors, such as provider-client interaction, and how they affect caregivers’ knowledge and exposure to information on Penta vaccination uptake has yet to receive sufficient attention in Nigeria. Previous studies ^22,24,25^ have shown that good interaction between the client and provider of care influences healthcare utilization and affects decision-making and acceptance of recommended health behaviour. Additionally, in terms of the impact of knowledge on child vaccination uptake, communities where vaccination enjoys a high level of acceptance and is made a social norm have been associated with increased vaccination uptake ^26–30^. Furthermore, exposure and adequate access to information can certainly enhance knowledge. For instance, a household survey in Sierra Leone found exposure to multiple sources of information about child vaccination was positively associated with child vaccination uptake. Specifically, exposure to information about child vaccination from religious leaders, health facilities, and community health workers was positively associated with child vaccination uptake ^23,31,32^. In general, studies ^29,33–36^ have explored the interaction between patient-provider interaction, caregivers’ exposure to child vaccination messages, knowledge about child vaccination, and vaccination uptake. However, there are limited studies in Nigeria and the northern region in particular that have explored caregivers’ first-hand experiences on the quality of counselling services received from providers regarding childhood vaccination and its implication for knowledge and vaccination uptake. Moreover, the coverage information was largely obtained from the child’s health card, thereby reducing the amount of possible recall errors from caregivers. Hence, this study examined the influence of quality counselling, caregivers’ exposure to vaccination messages, and uptake of Penta vaccine among caregivers of under-five children in Northern Nigeria. The choice of northern Nigeria is because it has the lowest coverage of all vaccination coverage including Penta vaccine relative to the southern region of Nigeria.

## MATERIALS AND METHODS

### Study Settings, Population, and Design

The study was conducted in six northern Nigeria states located in the northwest (Kano, Kaduna, and Sokoto) and northeast (Bauchi, Borno and Yobe) States (Fig. 1). These regions have the poorest health indices in the country including low child vaccination coverage and high childhood mortality^2^. The study population was all caregivers aged 15-49 years residing in the study area. We adopt a cross-sectional survey design to collect quantitative data from caregivers of children under the age of two years.

**Figure 1.**
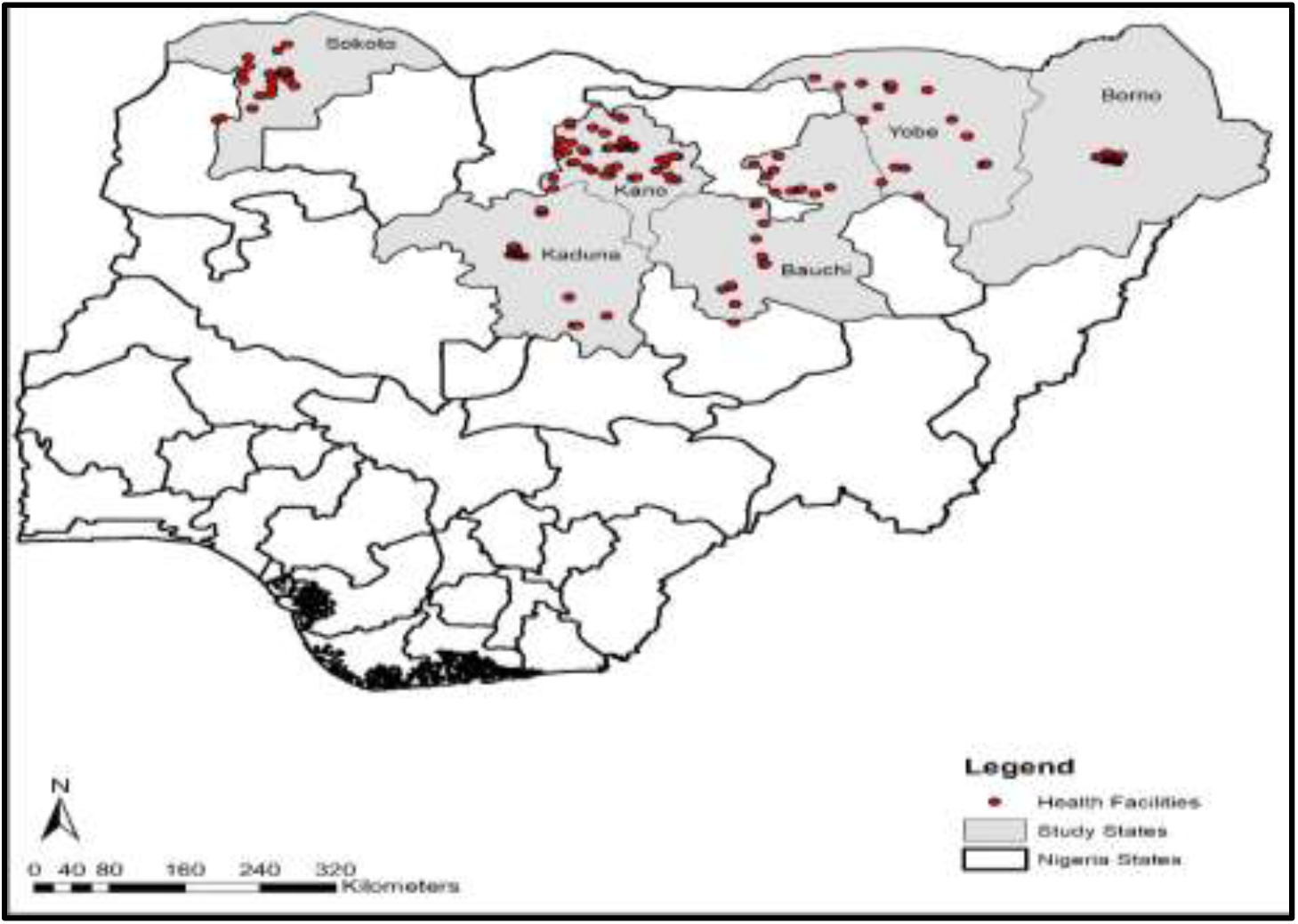
Map of Nigeria showing the study states.

### Sampling and Size

We used a two-stage stratified sampling procedure in selecting primary healthcare centers (PHCs). In the first stage, the states had three (3) senatorial districts each. To ensure coverage and spread, fifty percent of the total number of the Local Government Areas (LGAs) spread across the three senatorial districts in each state were selected proportionately (with the exception of Borno, where fewer LGAs were covered due to insecurity). For example, in a state with 20 LGAs half or 10 LGAs were randomly selected. In addition, two PHCs were randomly selected in each LGA, resulting in 20 health facilities sampled for that state. The same procedure was applied to other states except Borno.

A total of 1,077 exit interviews (CEIs) with caregivers accessing child vaccination services across all six states were conducted. However, for the findings presented in this study, the analysis was performed on 561 children aged 2-24 months who had received the Penta vaccine. The choice of Penta vaccine was because of its low coverage relative to other vaccines taken, while the age range 2-24 months was chosen because, at that age, the child is expected to have taken at least the first dose of Penta vaccine according to the national immunization schedule shown in Table 1. Additionally, the majority of the children were in the younger age group (< 6 months of age). Hence, the final sample size for this study is 561 caregivers of children aged 2-24 months who have been vaccinated with at least one dose of the Penta vaccine. According to the National Immunization schedule in Nigeria, Penta-1, 2, and 3 are expected to be taken at weeks 6, 10, and 14, respectively, at a dose of 0.05 ml administered at the left outer thigh of the child.^37^

**Table 1:** Explanatory Variables.

| Variable | Coding | Description |
| --- | --- | --- |
| Caregivers age | 15-24 years (1), 25-34 years (2), and 35-49 years (3) | Caregivers' age was collected as at the last birthday but categorized into three groups: |
| Level of education | No formal education (1), primary education (2), secondary education (3) and tertiary education (4). | Highest level of education attained |
| Occupation | Housewife (1), self-employed (2), teaching profession (3), student (4), and others comprising of civil servants and any other occupation category not captured (5). | Current occupation of the caregiver |
| Primary decision maker on child vaccination | Father (1), mother (2), and joint decision (3) | The primary decision maker on routine child vaccination |
| Child's sex | Male (1) or female (2) | Whether the child is male or female |
| Child's age | Integer recoded into 2-11 months (1) and 12-24 months (2). | Age as at last birthday |

### Inclusion Criteria and Sample Size

To be eligible for selection, caregivers must be aged 15-49 years with a child aged 0-24 months accessing child vaccination at the health facility at the time of the survey and willing to provide informed consent. Questions about child vaccination were asked with reference to the last child where the caregiver had more than one child. Caregivers were recruited at the point of presenting their child for vaccination because the visit to the health facilities was done on the days of the weekly routine immunization according to the schedule of the different health facilities.

### Data Collection

We conducted a client exit interview (CEI) with consenting eligible respondents between 14^th^ March and 8^th^ April 2022. All data gathering activities were held at the PHC facilities. The CEIs were programmed on mobile tablets using the SurveyCTO data collection platform. Several quality control measures were instituted to limit data entry errors and reduce cases of missing data. Additionally, the field team supervisors conducted daily reviews of data entry forms and reported on completeness, accuracy, timeliness, and inconsistencies before the data were finally uploaded to the server. The study tools were translated into Hausa (the predominant language in the study area) for meaning retention. Research assistants and supervisors had copies of the translated tools for possible reference during data collection to ensure that key terms were understood by respondents. The quantitative study tools were adapted from standard facility assessment questionnaires such as the WHO Service Availability and Readiness Assessment (SARA) Questionnaire.

## Measures

### Outcome Variable

The outcome variable is the uptake of any dose of the Penta vaccine, measured using a binary (coded “1” if received, otherwise “0”). The study controlled for some explanatory variables based on previous studies (Table 1).

At the multivariate level, the lowest value associated with a variable was used as the reference category. For instance, for caregiver age, the age group 15-24 years was used as the reference category. For all dichotomous variables, 0 was used as the reference category.

### Quality of Counselling (provider-client interaction)

Quality of counselling was measured using five indicators: whether the caregiver was told by the provider; what vaccine the child received; the date for the next vaccination appointment; the caregiver received information regarding possible side effects or reactions following the vaccination; the provider informed the caregiver on what to do in the event of a side effect; and whether the provider wrote down the date for the next vaccination appointment. To determine the quality of counselling provided, a composite score of all five indicators was generated. The categories in the good counselling (1) are those who reported yes to all five counselling indicators assessed; otherwise, counselling is considered poor (0).

### Caregivers’ Knowledge

To determine knowledge, caregivers were asked about when an infant should take the first vaccination dose after birth. Hence, women who reported that a child should receive their first vaccination immediately after birth or within the first week of birth were regarded as having correct knowledge, while those who reported later than one week were regarded as having incorrect knowledge.

### Awareness and level of exposure to child vaccination messages

Awareness was measured by asking if the caregiver had seen or heard of child vaccination in the last 30 days preceding the survey. The level of exposure on the other hand, was measured by the number of channels of information on child vaccination to which a caregiver is exposed. To avoid multicollinearity, only the level of exposure was used at the multivariate level. The level of exposure was determined using 13 different sources, including TV, radio, newspapers, health workers, CHWs, neighbors, community meetings, mother-to-mother support groups (MTMSG), other older children’s schools, places of worship, internet/social media, telephone messages, and community leaders. These were used to generate a composite score, while the mean and median values were determined. Thus, women with scores above the median value (in this case, heard messages from at least three different sources) were regarded as having good exposure (coded as 1); otherwise, they were regarded as poor (0).

## Statistical Analysis

First, descriptive statistics of the study variables, namely, frequency, count, and percentage, were performed. Secondly, in the multivariate model, we performed unadjusted binary logistic regression for each of the explanatory variables with the outcome variable to determine association at the bivariate level. For the model, all the explanatory variables were included. The associated odds ratio and 95% confidence intervals were reported. Variables with an associated p-value <0.05 were considered statistically significant. Analysis of data was performed using Stata 14 software.

## Ethical approval and informed consent

The study was approved by the Population Council Institutional Review Board (Protocol number 992). In Nigeria, the National Health Research Ethics Committee (approval number NHREC/01/01/2007-17/01/2022) and the States’ Health and Research Ethics Board also approved the study. Informed consent was obtained from each participant either through signatures or thumb prints for those unable to sign.

## RESULT

### Sociodemographic Characteristics of Respondents

Table 2 presents the sociodemographic characteristics of the study participants. The mean age of the caregivers was 27 years (median = 26 years) and ranged from 16 to 45 years. Nearly half (42%) of the women had no formal education. The mean age of the children was 7 months and ranged between 2 and 24 months.

**Table 2:**
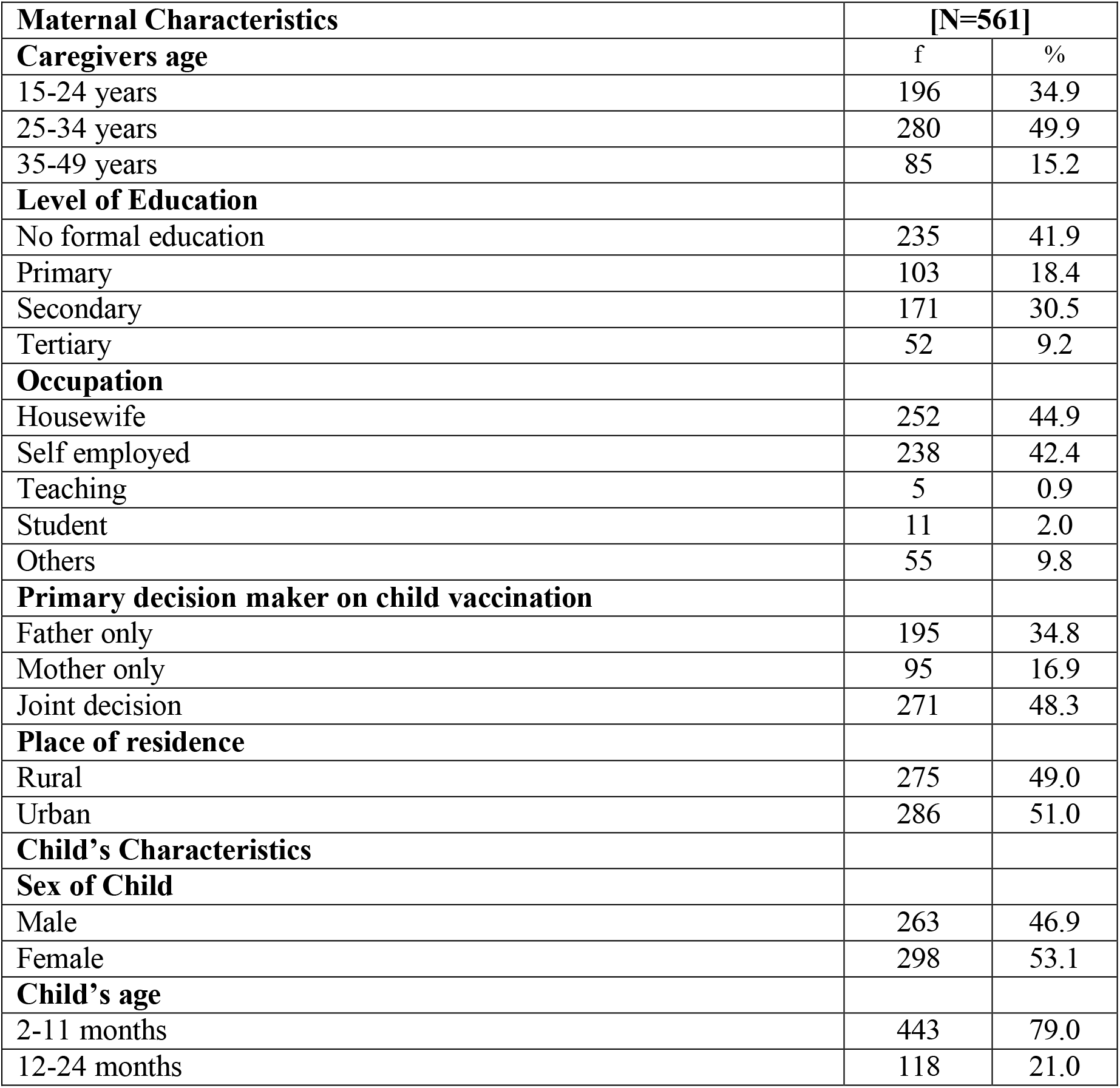
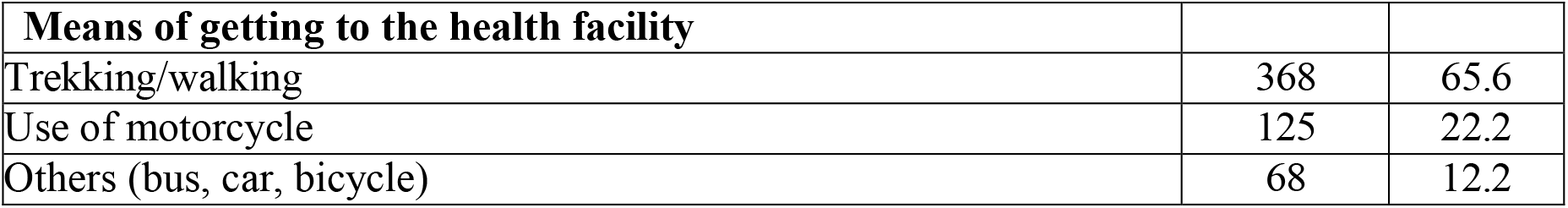
Sociodemographic Characteristics.

| **Maternal Characteristics** | **[N=561]** |  |
| --- | --- | --- |
| **Caregivers age** | f | % |
| 15-24 years | 196 | 34.9 |
| 25-34 years | 280 | 49.9 |
| 35-49 years | 85 | 15.2 |
| **Level of Education** |  |  |
| No formal education | 235 | 41.9 |
| Primary | 103 | 18.4 |
| Secondary | 171 | 30.5 |
| Tertiary | 52 | 9.2 |
| **Occupation** |  |  |
| Housewife | 252 | 44.9 |
| Self employed | 238 | 42.4 |
| Teaching | 5 | 0.9 |
| Student | 11 | 2.0 |
| Others | 55 | 9.8 |
| **Primary decision maker on child vaccination** |  |  |
| Father only | 195 | 34.8 |
| Mother only | 95 | 16.9 |
| Joint decision | 271 | 48.3 |
| **Place of residence** |  |  |
| Rural | 275 | 49.0 |
| Urban | 286 | 51.0 |
| **Child's Characteristics** |  |  |
| **Sex of Child** |  |  |
| Male | 263 | 46.9 |
| Female | 298 | 53.1 |
| **Child's age** |  |  |
| 2-11 months | 443 | 79.0 |
| 12-24 months | 118 | 21.0 |

| Means of getting to the health facility |  |  |
| --- | --- | --- |
| Trekking/walking | 368 | 65.6 |
| Use of motorcycle | 125 | 22.2 |
| Others (bus, car, bicycle) | 68 | 12.2 |

### Pentavalent Vaccine Coverage

The results shown in (Fig. 2) revealed that Penta vaccine coverage was low across all age groups. The coverage for Penta-3 was highest at 37% among children aged 12-24 months. Overall, only 43% of children aged 12-24 months received all three doses of the Penta vaccine.

**Figure 2.**
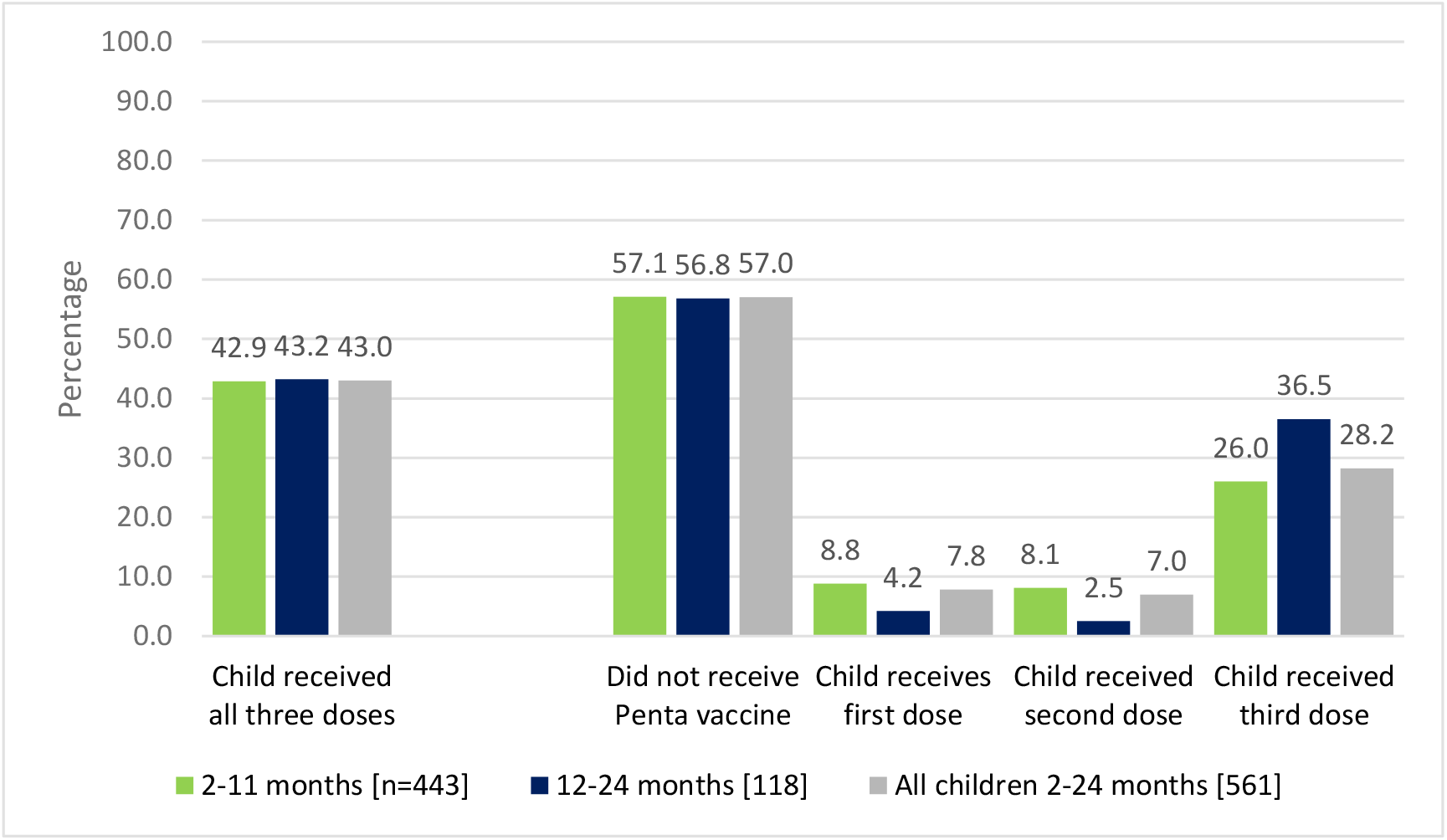
Pentavalent coverage according to doses.

### Quality of Child Vaccination Service, Caregivers’ Knowledge, and Exposure to Vaccination Messages

Findings revealed only 56% of caregivers received good counselling from the RI providers (Table 3). Concerning knowledge, a significant proportion (86%) correctly knew when a child should receive their first vaccination, with significant variations across doses. However, less than half (47%) correctly knew that a child should be vaccinated more than 5 times in their first year of life. Similarly, only 30% had good knowledge (comprising women who correctly mentioned a minimum of two purposes of child vaccination cards out of the three items assessed) of the purpose of vaccination cards.

**Table 3:** Child Immunization Service, Caregivers Knowledge, and exposure to vaccination messages.

|  |  | Penta Uptake |  |  |  |  |
| --- | --- | --- | --- | --- | --- | --- |
| Quality of provider-client interaction | Total n=561 | No Penta | first dose | second dose | Third dose | $\chi^2$/p value |
| Good | 56.1 | 56.5 | 8.6 | 5.7 | 29.2 | $\chi^2=2.381$ |
| Poor | 43.9 | 57.7 | 6.9 | 8.5 | 26.9 | p=0.497 |
| **Caregivers' Knowledge** |  |  |  |  |  |  |
| *Knowledge about the purpose of child vaccination card* |  |  |  |  |  |  |
| Good | 30.1 | 56.8 | 6.0 | 8.2 | 29.0 | $\chi^2=1.823$ |
| Poor | 69.9 | 57.1 | 8.7 | 6.4 | 27.8 | p=0.610 |
| *Knowledge about when a child should receive their first vaccination* |  |  |  |  |  |  |
| Correct | 86.0 | 54.1 | 7.9 | 7.9 | 30.1 | $\chi^2=13.776$ |
| Incorrect | 14.0 | 74.7 | 7.6 | 1.2 | 16.5 | p=0.003 |
| *knowledge about the number of vaccinations a child should receive in the first year of life* |  |  |  |  |  |  |
| Correct | 46.5 | 57.0 | 7.7 | 7.0 | 28.3 | $\chi^2=0.029$ |
| Incorrect | 53.5 | 57.0 | 8.0 | 7.0 | 28.0 | p=0.999 |
| *The caregiver had seen information on the child vaccination last 30 days* |  |  |  |  |  |  |
| Yes | 70.2 | 59.4 | 7.1 | 6.6 | 26.9 | $\chi^2=3.180$ |
| No | 29.8 | 51.5 | 9.6 | 7.8 | 31.1 | p=0.365 |
| *Level of Exposure to vaccination messages* |  |  |  |  |  |  |
| Good ($\geq 3$ sources) | 34.0 | 41.0 | 11.9 | 9.0 | 38.1 | $\chi^2=29.286$ |
| Poor ($< 3$ sources) | 66.0 | 68.9 | 4.6 | 5.4 | 21.1 | p=0.001 |

Regarding exposure to child vaccination messages, 70% have heard or seen messages on child vaccination in the last 30 days preceding the survey. However, only 34% had good exposure, representing caregivers who had seen information on child vaccination from at least three different sources (Table 3). The top three sources of child vaccination are health workers (67%), radio (41%), and community health workers (28%).

### Bivariate Association

At the bivariate level (Table 3), only correct knowledge of when a child should receive the first vaccination was significantly associated with the Penta vaccine update (x^2^=13.776, p=0.003).

Concerning exposure, good exposure (those exposed to at least three sources) to child vaccination messages was significantly associated with Penta vaccine uptake (x^2^=29.286, p=0.001). Similarly, receiving child vaccination messages from specific sources, namely, radio (x^2^=9.914, p=0.019), telephone (x^2^=10.284, p=0.016), community leader (x^2^=38.995, p=0.001), community health worker (x^2^=22.107, p=0.001), and place of worship (x^2^=18.786, p=0.001), were significantly associated with child vaccination uptake, while sources such as television, newspaper, internet, health worker and community meetings were not significant (Table not shown).

### Multivariate Result

The result of the adjusted binary logistic regression (Table 4) showed that knowledge of when a child should receive their first vaccination, sources of messages on child vaccination, primary decision maker on child vaccination, and region of residence were significantly (p<0.05) associated with the uptake of Penta vaccine.

**Table 4:** Binary logistic regression showing predictors of Penta uptake.

| Variables | Unadjusted Odds Ratio |  |  | Adjusted |  |  |
| --- | --- | --- | --- | --- | --- | --- |
|  | Odds Ratio | 95% Confidence Interval | p-value | Odds Ratio | 95% Confidence Interval | p-value |
| **Quality of counselling** |  |  |  |  |  |  |
| Poor | 1.000 |  |  | 1.000 |  |  |
| Good | 1.051 | 0.75-1.47 | 0.773 | 1.273 | 0.80-2.11 | 0.347 |
| **Caregivers' knowledge of when a child should receive the first vaccine** |  |  |  |  |  |  |
| Incorrect | 1.000 |  |  | 1.000 |  |  |
| Correct | 2.498 | 1.46-4.28 | 0.001 | 2.084 | 1.01-4.29 | 0.046 |
| **Exposure to vaccination messages** |  |  |  |  |  |  |
| Poor | 1.000 |  |  | 1.000 |  |  |
| Good | 3.174 | 2.06-4.89 | 0.001 | 0.922 | 0.42-2.02 | 0.840 |
| **Radio as a source of RI messages** |  |  |  |  |  |  |
| No | 1.000 |  |  | 1.000 |  |  |
| Yes | 1.854 | 1.23-2.79 | 0.001 | 0.969 | 0.54-1.73 | 0.914 |
| **Place of worship as a source of RI messages** |  |  |  |  |  |  |
| No | 1.000 |  |  | 1.000 |  |  |
| Yes | 3.953 | 1.83-8.52 | 0.001 | 2.777 | 1.15-6.67 | 0.022 |
| **CHW as a source of RI messages** |  |  |  |  |  |  |
| No | 1.000 |  |  | 1.000 |  |  |
| Yes | 2.583 | 1.65-4.04 | 0.001 | 1.949 | 1.14-3.34 | 0.015 |
| **Community leader as a source of RI messages** |  |  |  |  |  |  |
| No | 1.000 |  |  | 1.000 |  |  |
| Yes | 4.502 | 2.66-7.62 | 0.001 | 2.213 | 1.11-4.41 | 0.023 |
| **Neighbour as a source of RI messages** |  |  |  |  |  |  |
| No | 1.000 |  |  | 1.000 |  |  |
| Yes | 2.555 | 1.58-4.12 | 0.001 | 1.079 | 0.57-2.06 | 0.816 |
| **Primary decision maker on child vaccination** |  |  |  |  |  |  |
| Father | 1.000 |  |  | 1.000 |  |  |
| Mother | 0.456 | 0.26-0.79 | 0.005 | 0.319 | 0.15-0.67 | 0.002 |
| Joint decision | 1.310 | 0.91-1.91 | 0.142 | 1.030 | 0.62-1.71 | 0.907 |
| **Region of residence** |  |  |  |  |  |  |
| Northeast | 1.000 |  |  | 1.000 |  |  |
| Northwest | 1.349 | 0.96-1.89 | 0.083 | 2.601 | 1.51-4.48 | 0.001 |

Caregivers with correct knowledge of when a child should receive the first vaccination were twice as likely to receive the Penta vaccine (aOR=2.08; 95% CI=1.01-4.29) for their child.

Higher odds of Penta vaccine uptake were found among caregivers who received messages about child vaccination from a place of worship (aOR=2.78; 95% CI=1.15-6.67) and community leaders (aOR=2.21; 95% CI=1.11-4.41).

Receiving child vaccination messages from a Community Health Worker (CHW) was associated with nearly two times higher odds of a child receiving the Penta vaccine (aOR=1.95; 95% CI=1.14-3.34). On the other hand, lower odds of receiving the Penta vaccine were found when the mother was the primary decision maker on child vaccination (aOR=0.32; 95% CI=0.15-0.67).

Overall, children of caregivers residing in the northwest region were about three times (aOR=2.60; 95% CI=1.51-4.48) more likely to receive the Penta vaccine relative to their northeast counterparts.

## DISCUSSION

We assessed how the quality of counselling (provider-client interaction), caregivers’ knowledge, and exposure to vaccination messages affect the uptake of the Penta vaccine among children 2-24 months in six northern states in Nigeria. First, our study found low Penta vaccine coverage across all age groups, consistent with recent findings^13^. Previous studies have attributed the low coverage and low uptake of the first and second doses of the Penta vaccine to high dropout rates among infants who received the first but not the second dose^38^. These have been attributed to factors including lack of satisfaction with service and poor perception of benefits of vaccination among caregivers^3,4^. Similarly, another study investigating the determinants of uptake of a third dose of Penta found that children who received the first dose promptly were more likely to complete successive immunizations in accordance with the schedule ^39^. This suggests the need to encourage mothers to present their children early for the initial vaccination, in addition to addressing behavioural and institutional factors that could impede these caregivers from the timely presentation of their child for vaccination.

Our study also found that good knowledge regarding when a child should receive the first vaccination was significantly associated with the uptake of the Penta vaccine for the child. This positive association is very important considering the low prevalence of facility delivery and antenatal attendance among caregivers in the northern region of the country^13,40^. The likelihood of non-facility delivery among these women apart from negatively affecting knowledge can also affect the timely presentation of the child for vaccination^41^. Besides, it has been shown that adequate knowledge regarding vaccination was inversely associated with a delay in uptake^42^. In addition, our study found that receiving child vaccination messages from a place of worship, a community health worker, and traditional leaders was significantly associated with the uptake of the Penta vaccine. This is consistent with a recent study in Sierra Leone^31^ that found that exposure to information on child vaccination from religious leaders and community health workers was associated with child vaccination uptake. However, contrary to the Sierra Leone study, which found exposure to multiple sources of information to be positively associated with vaccination uptake, our present study found no significant association. The significant effect of traditional and religious leaders on child vaccination uptake is very important because of their positive influence in promoting child vaccination, especially in the northern region of the country. Over the years, traditional and religious leaders have proven to be instrumental in championing child vaccination and shaping norms and behaviour, particularly at the community level, where coverage is usually low^18^.

Finally, we found that low-quality counselling had no significant effect on the uptake of the Penta vaccine. Notwithstanding, the outcome of our study supports the ongoing effort toward strengthening interpersonal communications and interaction between health providers and their clients.^26^ This is critical, because of the importance of effective communication on healthcare delivery and quality. Quality counselling from providers to caregivers will no doubt help to build mutual trust and address bias, myths, and misconceptions about immunization, especially in the northern region of the country, where culture and traditions exert a strong influence on people’s way of life and acceptance of basic health interventions.

This study has some limitations regarding the sample in which the majority of the children were in the younger age group (below 6 months of age). Hence, due to the small sample size, there was very little allowance needed to restrict our analysis to the age group 11-23 months to better explore the extent of coverage. Additionally, the responses of the caregivers are largely based on current experience and might not be a true reflection of things. Furthermore, the findings only capture the experiences of caregivers within the health facility, excluding the experiences of women in the community. Notwithstanding these limitations, our present study has made an important contribution to the literature by stressing the importance of counselling and the role of traditional and religious institutions in promoting improved child vaccination coverage.

## CONCLUSION

Effective communication strategies are critical to immunization uptake among caregivers because they can enhance their knowledge about the benefits of immunization and correct false impressions or concerns that often prevent caregivers from getting their children immunized. Hence, our findings suggest the need for continuous strengthening of the capacity of routine immunization health workers, by providing them with requisite skills and strategies to interact with their clients. This will build trust, a very important component for vaccination uptake, and address issues around vaccine hesitancy. This study further highlights the important role of community and religious institutions in championing routine child vaccination, especially in the study areas. This is an important consideration for policy makers, program implementers, and relevant stakeholders working towards improving routine child vaccination coverage in Nigeria.

## LIMITATION OF THE STUDY

One major limitation of this study is that the majority of the children were in the younger age group (<6 months of age) and therefore did not give much room for restricting our analysis to the age group 11-23 months to better explore the extent of coverage due to sample size. While sampling was done randomly in most cases at the health facilities, there were fewer instances where caregivers who met the inclusion criteria were found at the health facility. In these cases, all the women were sampled. This might have introduced potential bias. Also, the responses of the caregivers are largely based on current experience and might therefore not be a true reflection of things. Furthermore, findings only capture the experiences of caregivers within the health facility, excluding the experiences of women in the community. More so, the use of cross-sectional survey data poses some limitations concerning establishing causal relationships. Notwithstanding these limitations, our present study has made an important contribution to the literature.

## Funding

This research article is based on research supported by the Gates Foundation with funding code [INV-033024] implemented in six northern Nigerian states of Bauchi, Borno, Kaduna, Kano, Sokoto and Yobe states. Also, the findings and conclusions arrived at reflect those of the authors and do not in any way reflect the positions of the Gates Foundation.

## Data Availability Statement

All data relevant to the publication can be obtained upon reasonable request to the Population Council. Contact Matthew Alabi at.

## Conflict of Interest

The authors declare none

## Notes

### Competing Interest Statement

The authors have declared no competing interest.

### Author Declarations

The study was approved by the Population Council Institutional Review Board (Protocol number 992). In Nigeria, the National Health Research Ethics Committee (approval number NHREC/01/01/2007-17/01/2022) and the States Health and Research Ethics Board also approved the study.

